# Simulating Pandemic Disease Spread and the Impact of Interventions in Complex Societal Networks

**DOI:** 10.1101/2020.10.28.20221820

**Authors:** Daniel S. Mytelka

## Abstract

**Introduction:** Projecting disease spread is challenging because of the heterogeneous nature of human interactions, including both natural societal interactions and how they change in response to pandemics. Simulations can provide important guidance regarding the likely impact of interventions on an assumed base case.

**Methods:** This paper uses assumptions based on the COVID-19 pandemic to construct a Susceptible-Infectious-Recovered model representative of US society, focusing on the interrelationships of groups with differing contact networks (essential/non-essential workers and urban/non-urban populations). The model is used to explore the impact of interventions (reduced interactions, vaccinations and selective isolation) on overall and group-specific disease spread.

**Results:** In the absence of herd immunity, temporary interventions will only reduce the overall number of disease cases moderately and spread them over a greater period of time unless they virtually eliminate disease and no new infections occur from exogenous sources. Vaccinations can provide stronger benefit, but can be limited by efficacy and utilization rates.

**Conclusions:** While a highly effective and broadly utilized vaccine might halt disease spread, some combination of increased long-term surveillance and selective isolation of the most vulnerable populations might be necessary to minimize morbidity and mortality if only moderately effective vaccines are available.

## Introduction

During 2020, the novel coronavirus SARS-CoV-2, the etiological agent for COVID-19, spread from initial detection in Wuhan, China to virtually every region of the world ^1^. A variety of efforts have been made to model disease trajectory in the United States ^2^, but the resulting models are challenged by substantial uncertainties with regard to disease parameters, inconsistent official policy and the unpredictability of human behavior. While these factors may make absolute prediction of disease spread difficult, understanding parameters that influence disease spread and the relative impact of interventions can provide important qualitative public health learnings. The goal of this paper is to explore factors that influence or mitigate airborne disease spread using a baseline model that generally reflects current assumptions about COVID-19.

Modeling the spread of any airborne disease typically relies heavily on R_0_, the disease’s basic reproductive number. R_0_ reflects the average number of new cases that arise from every infected individual ^3^. If R_0_ is greater than 1, the disease will be spreading exponentially (more newly infected individuals in each infected generation), while if R_0_ is less than 1, the spread will be slowing. If individuals who survive an infection are immune to reinfection or individuals can be rendered immune through vaccination, the effective R_0_ for the disease will fall over time ^4^, providing a natural barrier to further spread of disease (herd immunity). Initial R_0_ for COVID-19 was estimated to be about 2.5 ^5^; other early estimates ranged from about 2 to more than 6, with most falling between 2 and 4 ^6^.

R_0_ may vary because of a number of factors, including mechanics of transmission, length of infectivity, magnitude of infectivity relative to symptoms, heterogeneity in number of contacts, likelihood that a contact leads to disease transmission, underlying nature of human contacts and disease control mechanisms. Importantly, human society is not structured such that individuals have equal chances of contacting any other individuals. Most contacts (particularly close and prolonged contacts) are with close friends and family, leading to increased odds of transmission within such close core groups ^7^. Highly infectious “super-spreaders” may also play a significant role in disease transmission ^8^. Some individuals may be relatively isolated; if they have few close contacts and those contacts either never become infected or fail to transmit disease when infected, they may remain disease naïve. Herrmann et al. ^9^ have shown the impact of modeling societies as networks rather than homogeneous groups with relatively random contacts and Britton et al. ^10^ have shown that considering societal subgroups can influence requirements for herd immunity.

Absent interventions, pandemics can be divided into three phases for modeling purposes: A ramp-up phase where the initially infected individuals may be few and localized, an exponential phase where the number of infected individuals is increasing rapidly and infection is extensive, and a post-exponential phase where the number of cases is declining. If a disease spreads poorly (low R_0_) or early infected individuals are isolated/fail to spread disease, disease spread may never become extensive.

This paper will start by exploring how connectivity impacts the spread of disease through a population, use this understanding to build a model that reasonably reflects the US population, and then look at the impact of potential interventions in this model. While no claims are made that the model accurately reflects COVID-19 spread, assumptions were chosen based on general current knowledge of this disease. General learnings are important with regard to current public policy choices on this disease, such as the controversy regarding whether a policy of accepting disease spread among those less likely to have severe disease consequences in order to develop some herd immunity and protect the most vulnerable is a good idea (Great Barrington Declaration^11^) or not (John Snow Memorandum^12, 13^).

## Methods

Modeling was conducted using Microsoft Excel Professional Plus 2016.

Simulations used Visual Basic. Unless otherwise specified, 100,000 simulations were performed for each example using a sample population of 1000 individuals, of whom one random individual was initially infected. In most scenarios, additional infections from exogenous sources were permitted.

For the purposes of this simulation, a simple Susceptible-Infectious-Recovered (SIR)-type model was used for viral transmission. Time was divided into periods intended to represent the primary infectivity period of individuals. During each period, currently infected individuals had disease-spreading contacts with a number of individuals pulled from a Poisson distribution that varied based on the infected individual’s characteristics. If the other individuals were susceptible, they would become infected and be part of the next cohort of infected individuals. Disease-spreading contacts were most likely to be with close members of the infected individual’s network, reflecting both the higher number and greater intensity of those interactions. Individuals were assumed to be infectious for only one period. The primary infectivity period might proxy a real-world period encompassing late asymptomatic and early symptomatic phases (perhaps five to nine days for COVID-19 in the real world ^14, 15^), and simulations were allowed to run for up to 146 periods (∼2^+^ years).

Note that in a population with homogenous individuals and random association, the population-level parameter R_0_ and the number of disease-spreading contacts by an individual are the same, but for heterogeneous populations, R_0_ will reflect a blend of the properties of individuals. As the number of previously infected individuals in the population rises, disease-spreading encounters are increasingly likely to be with immune individuals, leading to a reduction in individual-level disease spread paralleling the population-level shift from an initial R_0_ to effective R_0_.

Parameters that varied by simulation included the average number of infectious events, likelihood of contact between any two individuals, and likelihood of additional infections from outside the system. Factors could also vary between differently situated individuals within a scenario or over time (interventions). For example, one individual might be more isolated and have a smaller contact network and fewer infectious encounters, or might self-isolate after a pandemic starts.

For the examples using a representation of the US society, individuals were placed into a hierarchy that comprised a “core” group representing their most frequent contacts (household), a broader group of less frequent contacts that formed a subset of their class (e.g. urban essential workers), their class overall, and overall society. Parameters were chosen such that all individuals had similar likelihoods of transmitting disease to members of their closest “core” group (household), but members of different classes had different likelihoods of interacting outside of their core group (See Table 1). Urban individuals were assumed to have more contacts overall and essential workers were assumed to have both more contacts and more contacts with individuals outside of their close network. Assumptions were roughly adjusted to give an early disease spread rate similar to what was observed for COVID-19 in the US.

**Table 1.** Simulation parameters representative of US society (See Methods for rationales).

| % Urban | 31% | Parker et al.<sup>28</sup> |  |  |
| --- | --- | --- | --- | --- |
| Essential | 55M | McNicholas and Poydock<sup>29</sup> |  |  |
|  | Urban<br>Non-Essential | Urban<br>Essential | Non-Urban<br>Not-Essential | Non-Urban<br>Non-Essential |
| N | 250 | 50 | 550 | 150 |
| Core Interaction Group | 5 | 5 | 5 | 5 |
| Broader Interaction Group | 25 | 25 | 10 | 25 |
| Average Infectious Contacts | 4 | 6 | 2.5 | 4 |
| % to Core | 50% | 33% | 80% | 50% |
| % to Broader | 40% | 47% | 10% | 30% |
| % to others in class | 8% | 16% | 8% | 16% |
| % other classes | 2% | 4% | 2% | 4% |
| Exogenous Infections | 50% (chance of one new infection per period) |  |  |  |

Limited disease spread is defined as situations where disease does not spread broadly through the population despite parameters that could support broad spread. Disease spread may be limited due to chance (early infected persons do not spread disease broadly) or the nature of the societal connections (individuals serve as bottlenecks). In simulations, cases where disease spread is 10% or less of the initial population were characterized as limited (as opposed to extensive) disease spread.

For the analysis of time to elimination of disease from a cohort through isolation alone, a simple cohort model with a homogeneous population was used wherein each individual had an identical probability of infecting one individual in the next period. The initial cohort had 250,000 infected individuals and results were the average of 1000 simulations. There were no infections from exogenous sources and the population size was deemed sufficient large such that the number of previously infected (and presumptively immune) individuals was not relevant.

## Results

### Designing an Appropriate Model to Represent US Society

In a simple homogeneous model where individuals have equal chances of spreading disease to any other individuals, disease spread is based primarily on the average number of contacts an individual has that transmit disease (“infectious contacts”), a proxy for the population-level R_0_ (See Methods). If early cases have few infectious contacts, disease spread may remain limited, but if disease spreads beyond a few individuals or new cases continue to arise from exogenous sources, disease spread will generally be extensive given a sufficiently high average number of infectious contacts (data not shown).

From a modeling perspective, situations where initial disease is not contained are the relevant public health scenarios. The pace of disease spread and average number ultimately infected increase with the average number of infectious contacts (See Figure 1). Over time, the number of susceptible individuals in the population declines (assuming that previously infected individuals are resistant to reinfection) and an increasing proportion of contacts that would otherwise spread disease can no longer do so (matching the population-level decline in effective R_0_). The length of time until peak increases with the log of the population size (data not shown).

**Figure 1.**
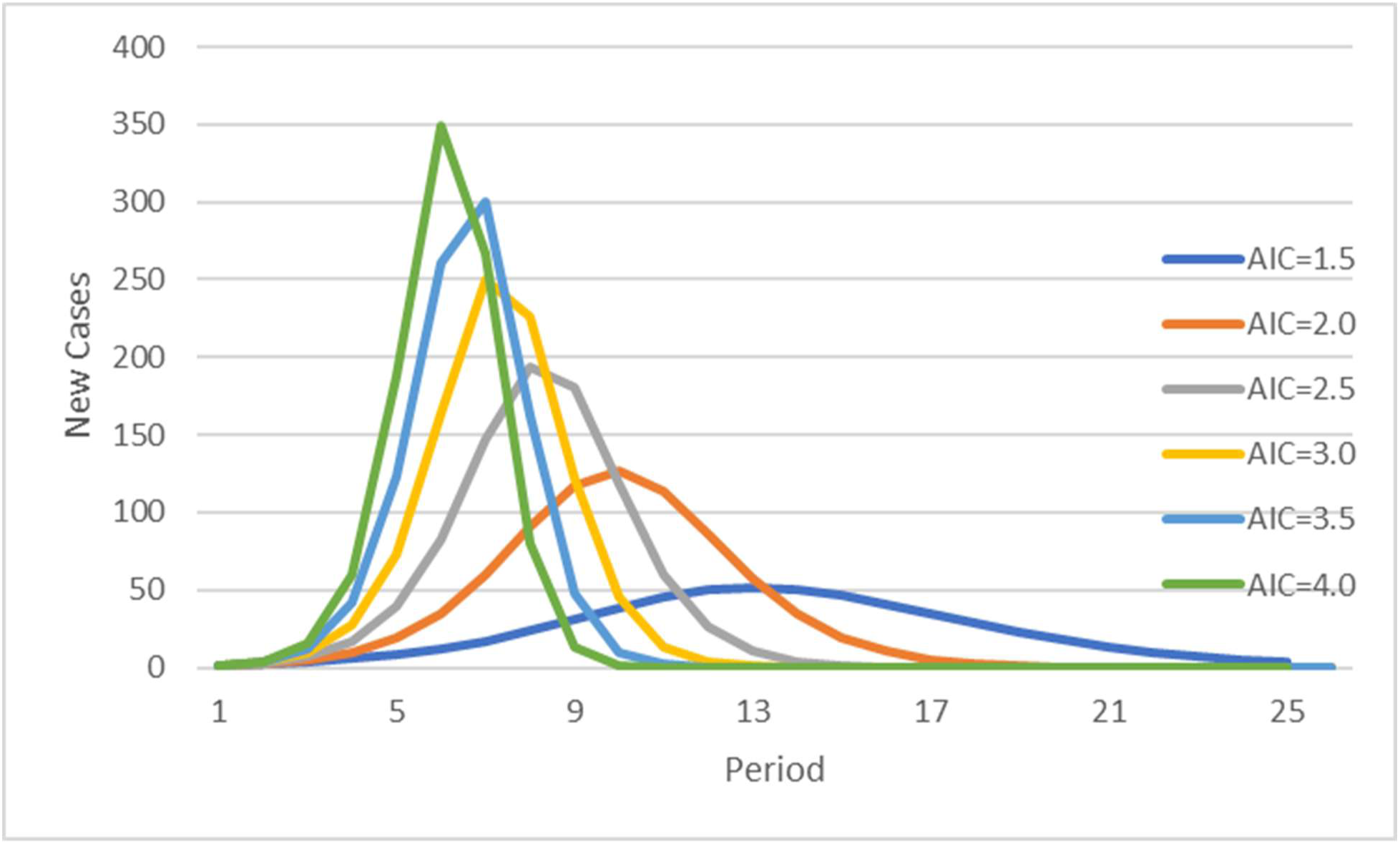
Differences in disease spread with average infectious contacts (AIC). Each curve represents the results of 100,000 simulations of 1000 homogeneous individuals with the specified number of average infectious contacts (the individual-level proxy for R_0_). Only simulations with extensive disease spread (>=10% of overall population) were included, representing 57%, 79%, 90%, 94%, 97% and 98% of simulations respectively for AIC of 1.5-4. Average number of individuals ultimately infected were 582, 796, 893, 940, 966 and 980 respectively.

There are multiple ways to model more heterogeneous individual contacts in real societies. Most contacts are likely to be with a small group of close family and friends, with a broader set of secondary contacts that varies by individual. One could model the core groups as relatively isolated, with only a small number of individuals having significant contacts outside of the inner group (See Figure 2A), core groups that have common external contacts (Figure 2B) or core groups where individuals have diverse external contacts (Figure 2C). Each of these structures has analogs in real societies: Elderly individuals with few contacts outside the house, common secondary associations with social groups (e.g. churches) or those geographically proximal, and differing places of employment or schools for different family members. Models where specific individuals are key intermediaries (Figure 2A) lead to substantial bottlenecks that limit spread of the disease when core connecting individuals either do not become infected or do not happen to infect individuals in other groups (data not shown), while models with very broad contact networks if not substantially constrained become similar to societies with homogeneous contacts. This paper uses a hybrid three-level model of society, wherein individuals have most of their contacts with a small group, a smaller number of contacts with a broader shared group, and a still smaller number of contacts with any other members of society.

**Figure 2.**
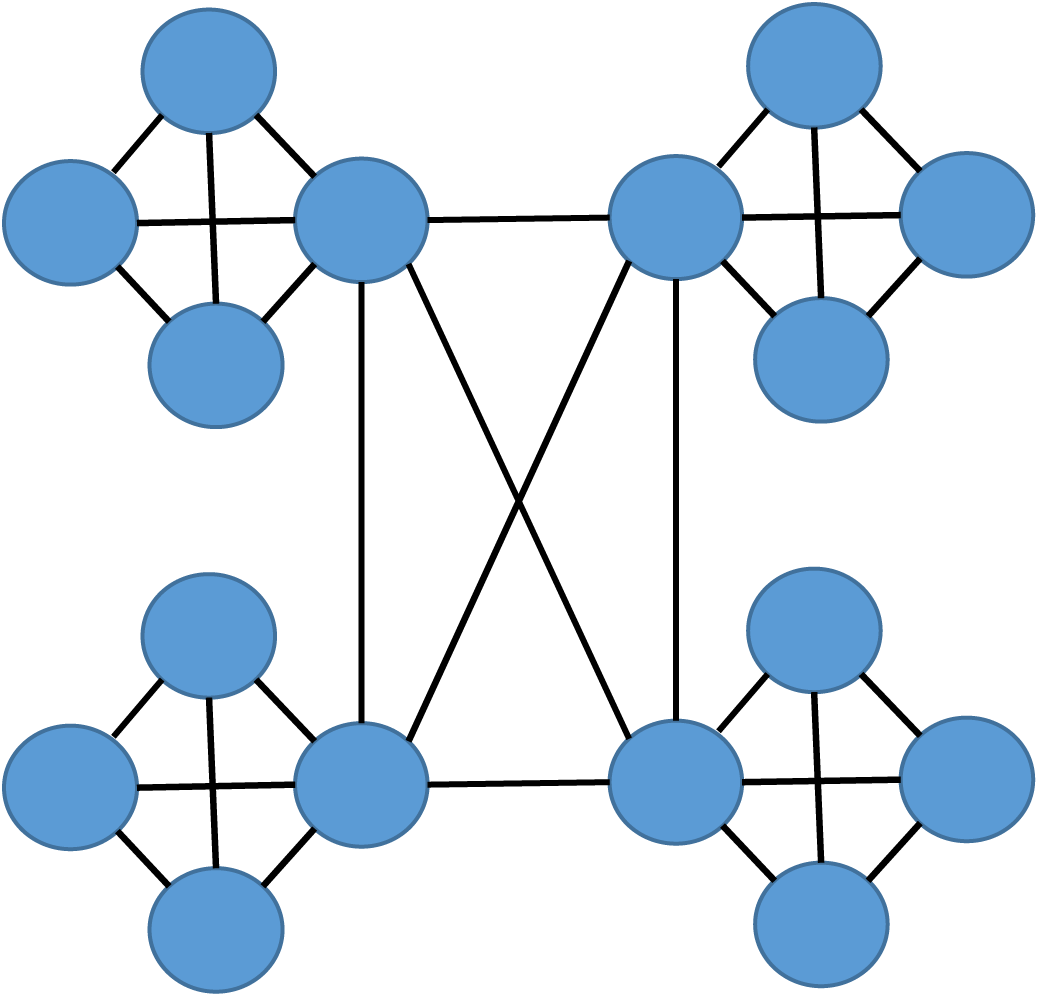
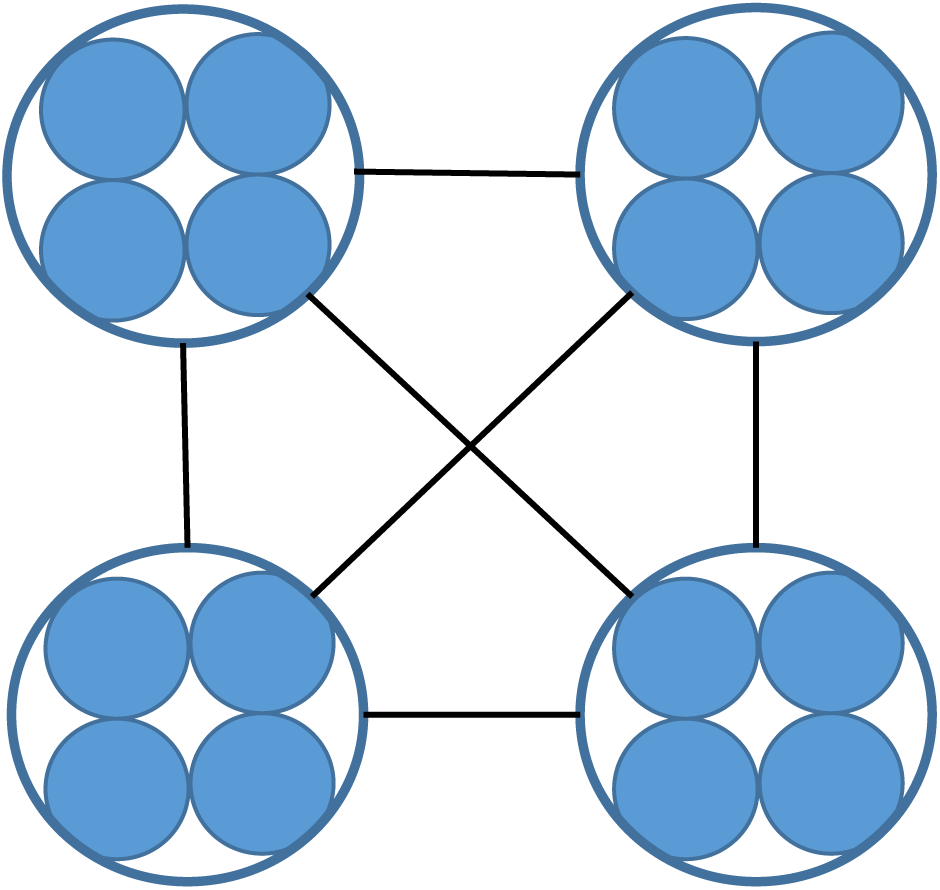
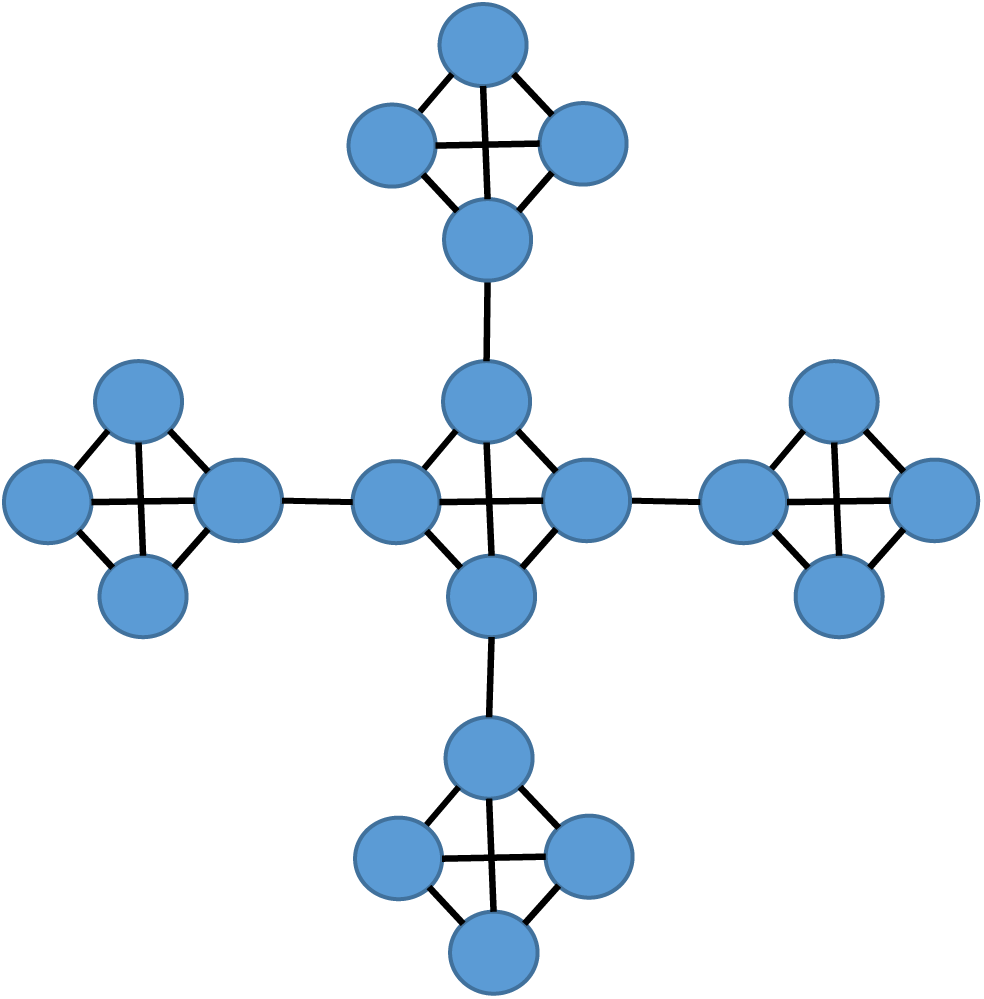
Potential models for interactions between members of a core interaction group (e.g. a family) and the broader community. A) Most individuals have few external contacts with one individual serving as the primary link to the external community. B) Individuals have most of their contacts within the core interaction group, and external interactions are similar among all members of the group. C) Individuals have most of their contact within the core interaction group, but external interactions differ by group member.

As the US society is extremely heterogeneous, it is challenging to construct a model that represents all aspects of society while remaining manageable and providing clear learnings with regard to how intercessions might impact it. However, a relatively simple model can prove informative. Two factors were chosen as most important for modelling: Population density and breadth of interactions. Generally, it appears that spread of disease is correlated with population density (See Appendix 1). While urban individuals do not have dramatically larger core contact groups based on average household sizes ^16, 17^, there are more opportunities to interact outside of the core group (e.g. apartment common areas, street crowds). Breadth of interactions reflects individual proclivities, but was significantly reduced in many individuals other than essential workers after the start of the pandemic. Table 1 shows parameters that were chosen for the base model before any interventions (See Methods for details), recognizing that some of these parameters were relatively arbitrary and chosen to make the model resemble real-world experience; a replication with a different set of parameters for sensitivity yielded virtually identical qualitative results to those shown below (data not shown).

In the absence of any behavioral changes to limit the spread of disease, this set of parameters leads to rapid spread of disease through the population, with urban classes typically hit faster and harder because of their larger typical number of contacts per individual and broader contact groupings (See Figure 3), despite random initiation of the disease that tends to be outside of urban areas due to the overall population distribution. Because of greater relative isolation of individuals, spread through non-urban areas also tends to be less complete, with 60% of the non-urban/non-essential population ultimately affected compared to at least 96% of the other three classes. Note that in the US as a whole, overall population is approximately 2^18^ times larger than the simulated population (1000), leading to a substantial increase in the time spent in each stage in a real population.

**Figure 3.**
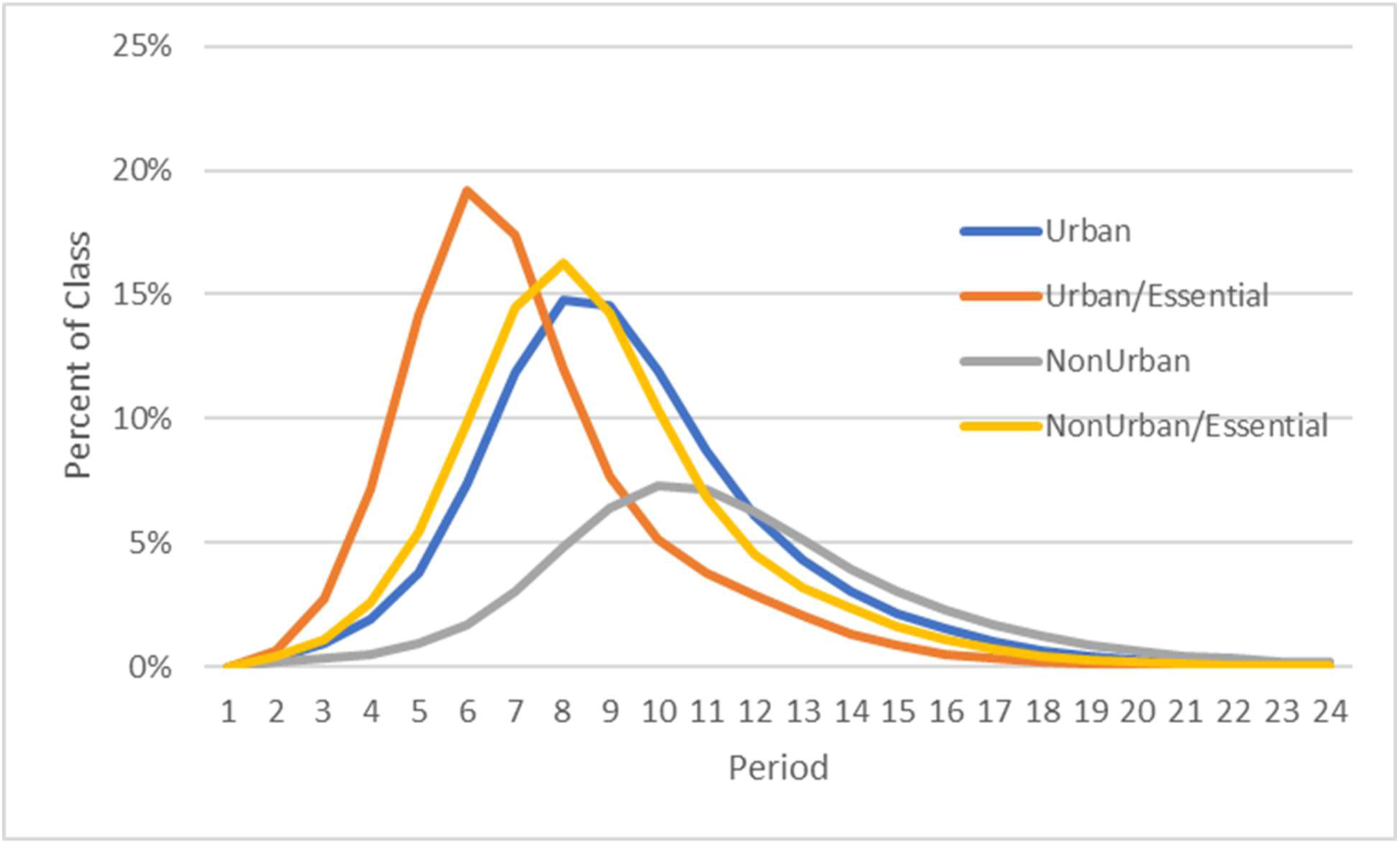
Spread of disease through different segments of the population. Simulations used the parameters described in Table 1, with initiation of disease in a random individual for each of the 100,000 simulations of 1000 individuals.

### Interventions to Limit Disease Spread

Interventions can affect disease spread through a population in four ways. First, they can temporarily reduce interactions to minimize current disease spread and the number of near-term affected individuals (“flatten the curve”). Second, they can make more permanent changes in interaction patterns that reduce overall speed of spread, such as tracking disease, reducing contacts and decreasing incentives for working sick. Third, they can decrease the number of people who are susceptible to disease, reducing both near-term and long-term infection (e.g. by vaccination). Finally, they can try to impact which individuals are infected, such as by making special provisions for those who are most likely to have serious disease consequences.

Figure 4 shows the impact of a temporary reduction in interactions. Interactions were virtually eliminated for non-essential workers from period 6-12, and reduced for essential workers. When restrictions were removed, the disease spread resumed because residual cases had remained in the population, recapitulating the initial ramp-up to exponential phases of disease spread, though cases were likely to be scattered and thus more challenging to control. More stringent or longer controls had greater opportunity to eliminate all cases in the absence of new infections from exogenous sources (Figure 5). After the interruption, the urban/essential group had substantially reduced number of cases due to many having already been infected in the first peak, helping to mitigate the overall spread across all groups by about 11%.

**Figure 4.**
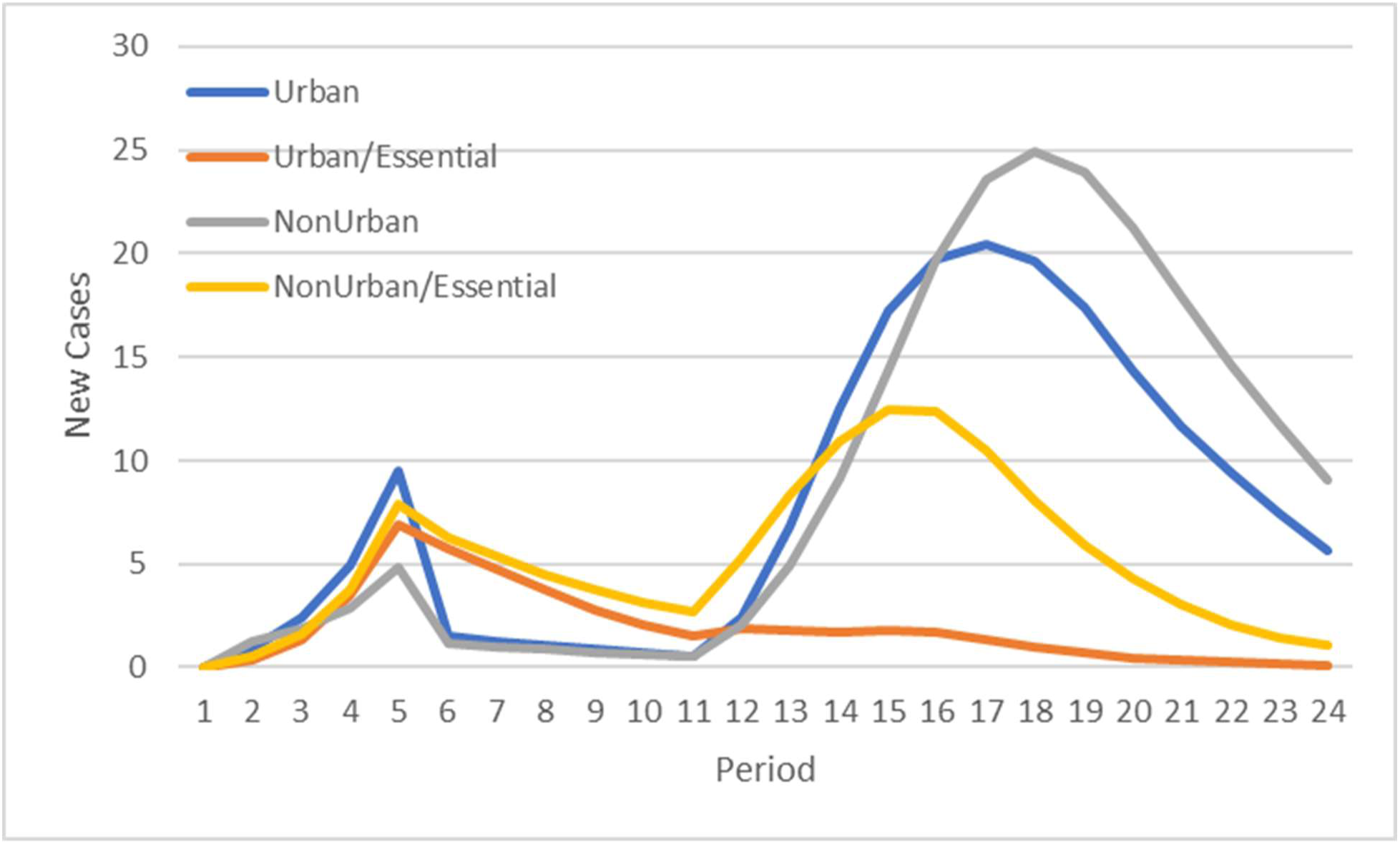
Spread of disease through the population with reduced interactions from period 4 to period 11 (80% reduction in essential classes and 99% reduction in other classes). Exogenous sources of infection were eliminated after period 4.

**Figure 5.**
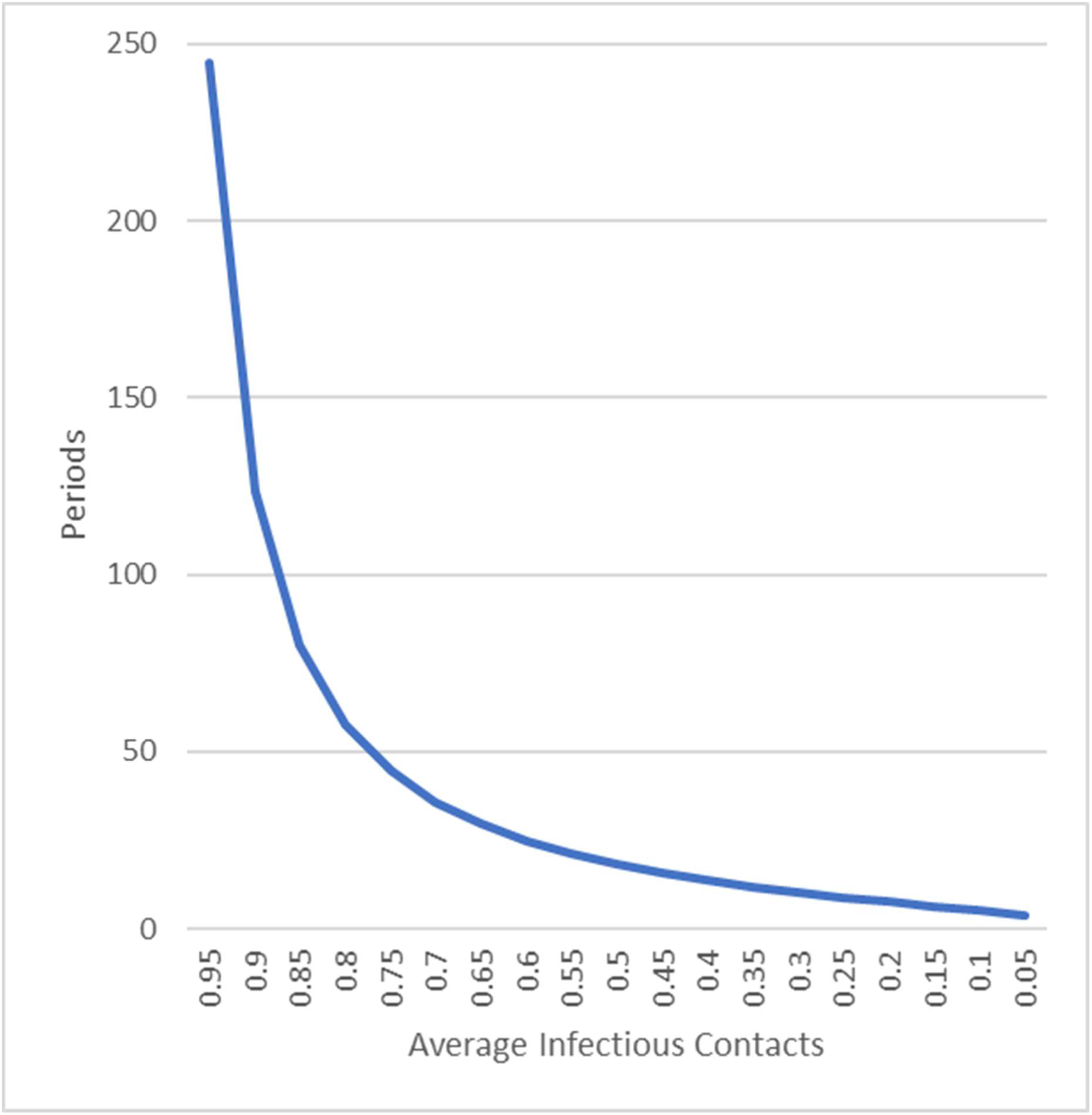
Time to elimination of disease through social distancing alone. Based on a simple cohort model (See Methods), 250,000 current cases of disease can be reduced to zero through draconian reductions in interactions, but an exponentially increasing number of periods are necessary if the decrease in contacts is not extreme.

Permanent changes in interaction patterns are likely to be more modest in nature, such as reductions in sick individuals outside of the house ^18^ and increases in working from home. These could reduce disease transmission in general, lowering R_0_, but likely not significantly enough to have major impact on COVID-19 except in combination with other control methods (See Figure 1). Aggressive use of testing and isolation of sick individuals could be used to control subsequent spread after a temporary reduction in cases down to a manageable level.

Vaccines are unlikely to be 100% effective. The FDA has proposed a 50% minimum point estimate of effectiveness for approval ^19^, in line with the 40-60% effectiveness seen for influenza vaccines ^20^. In addition, surveys suggest that 20-50% of individuals might refuse vaccination ^21^, with as many as 67% likely to delay vaccination ^22^. Figure 6 shows the reduction in benefit from vaccines that would result from less than complete coverage due to some combination of unvaccinated or ineffectively vaccinated individuals (but ignoring the loss due to delays in vaccination). Any reductions in susceptible individuals due to prior exposure would be synergistic with the vaccine benefit.

**Figure 6.**
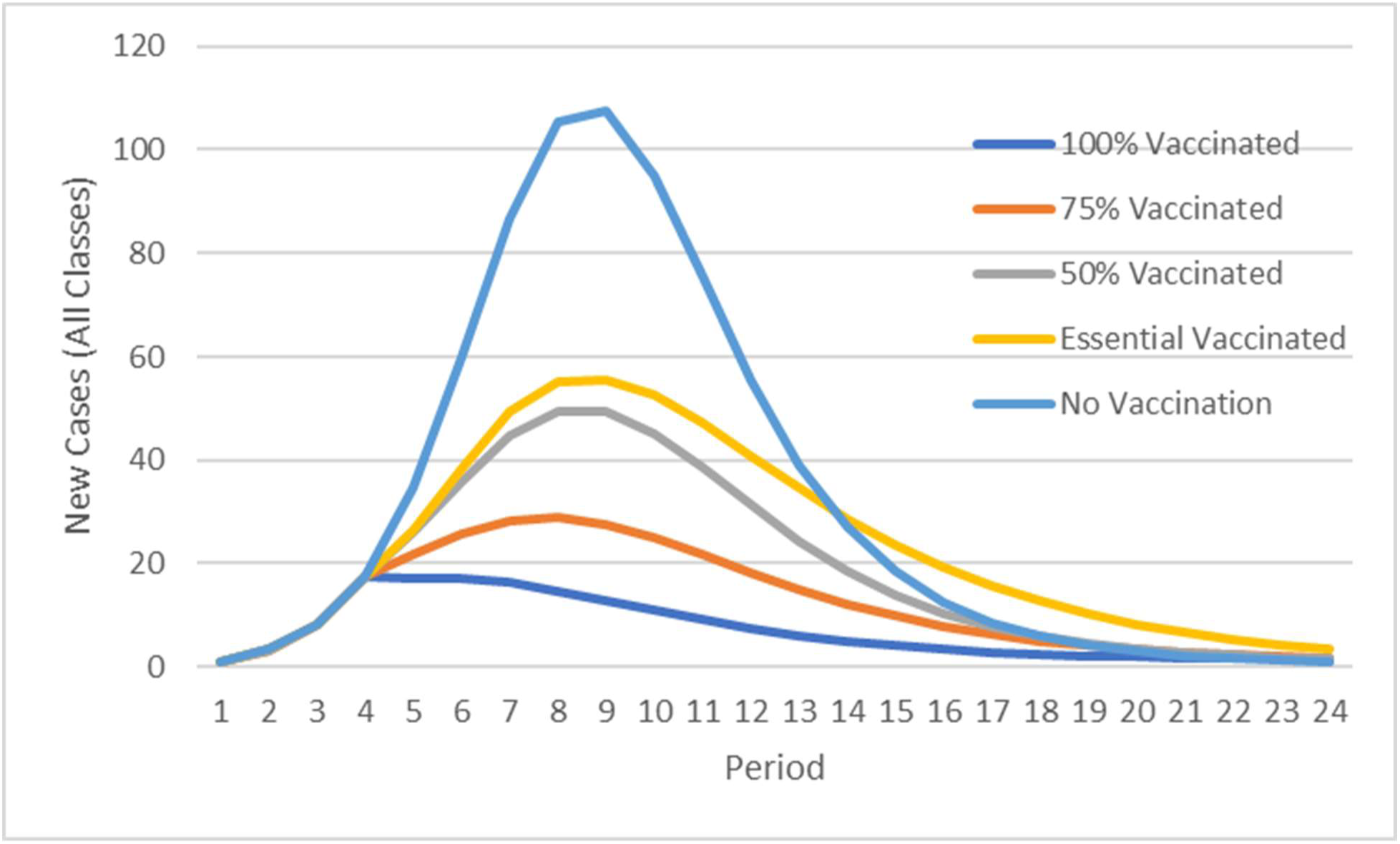
Spread of disease through the population with vaccination broadly administrated during period 4, showing the sum of new cases across all groups. Relative to no vaccination, the total number of infected individuals declines subsequent to period 4 by 19% (essential workers only vaccinated with 100% efficacy), 40% (50% effective vaccination), 58% (75% effective vaccination), and 74% (100% effective vaccination).

When only vulnerable groups such as older and less healthy individuals are isolated, infections will occur in other segments of the population during initial exponential disease spread and later provide some herd immunity for the vulnerable population. Figure 7 shows the impact of taking populations with initially similar transmission patterns (Figure 7A) and sequestering 25% of the population for various periods of time. The curve for the less vulnerable population shifts because there are assumed to be fewer interactions overall rather than a replacement of some interactions that would otherwise have occurred with the sequestered population. If the population that is sequestered emerges before the disease has run its course, there will be a resurgence of disease in both populations (Figure 7B), leading to a modest benefit of 19% and 3% reduction in infections in the vulnerable and less vulnerable groups respectively. However, if the vulnerable population remains sequestered until the disease has run through most of the less vulnerable segment of the population, the less vulnerable population will provide herd immunity for the more vulnerable population and reduce spread in the two groups by 60% and 14% (Figure 7C).

**Figure 7.**
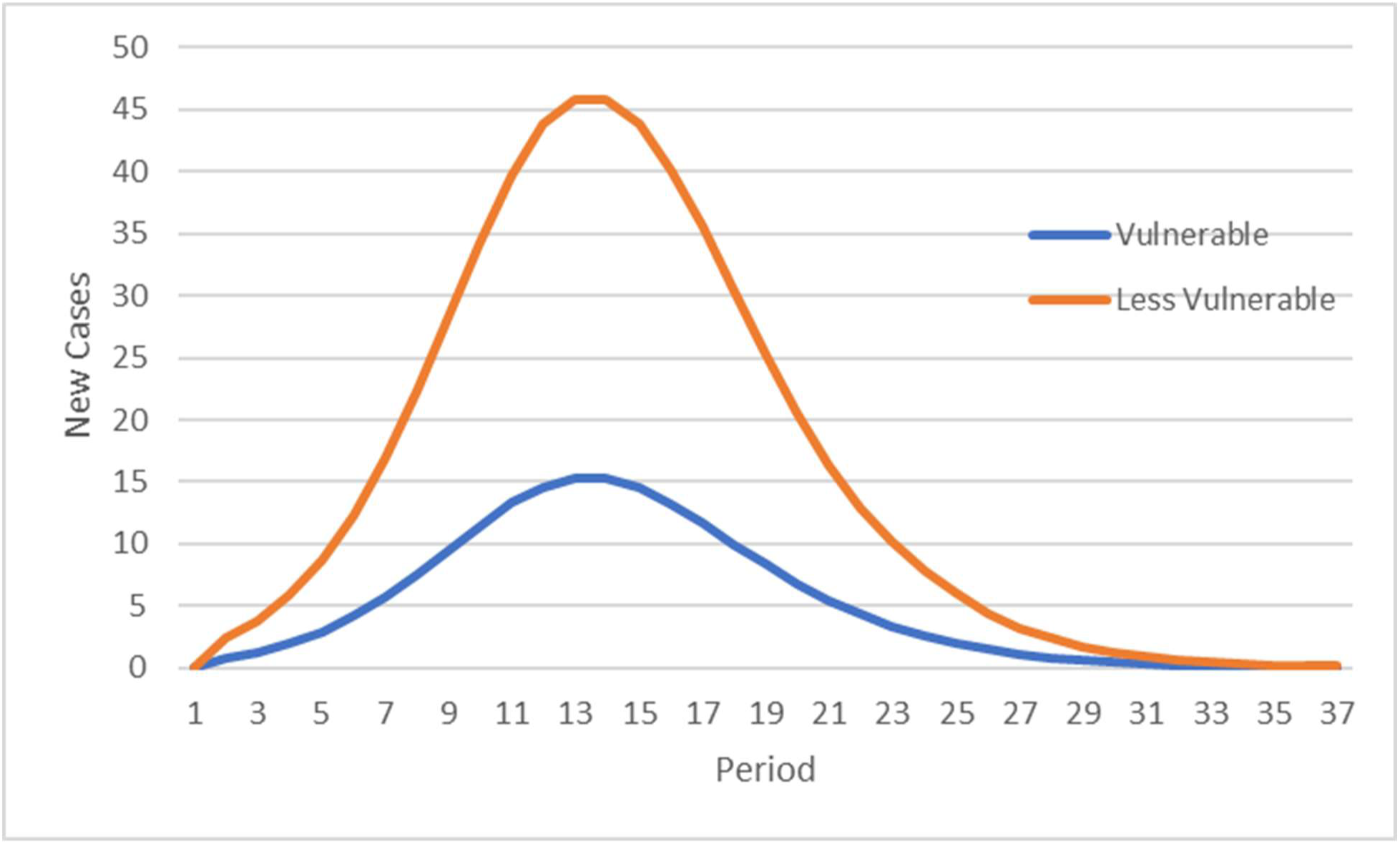
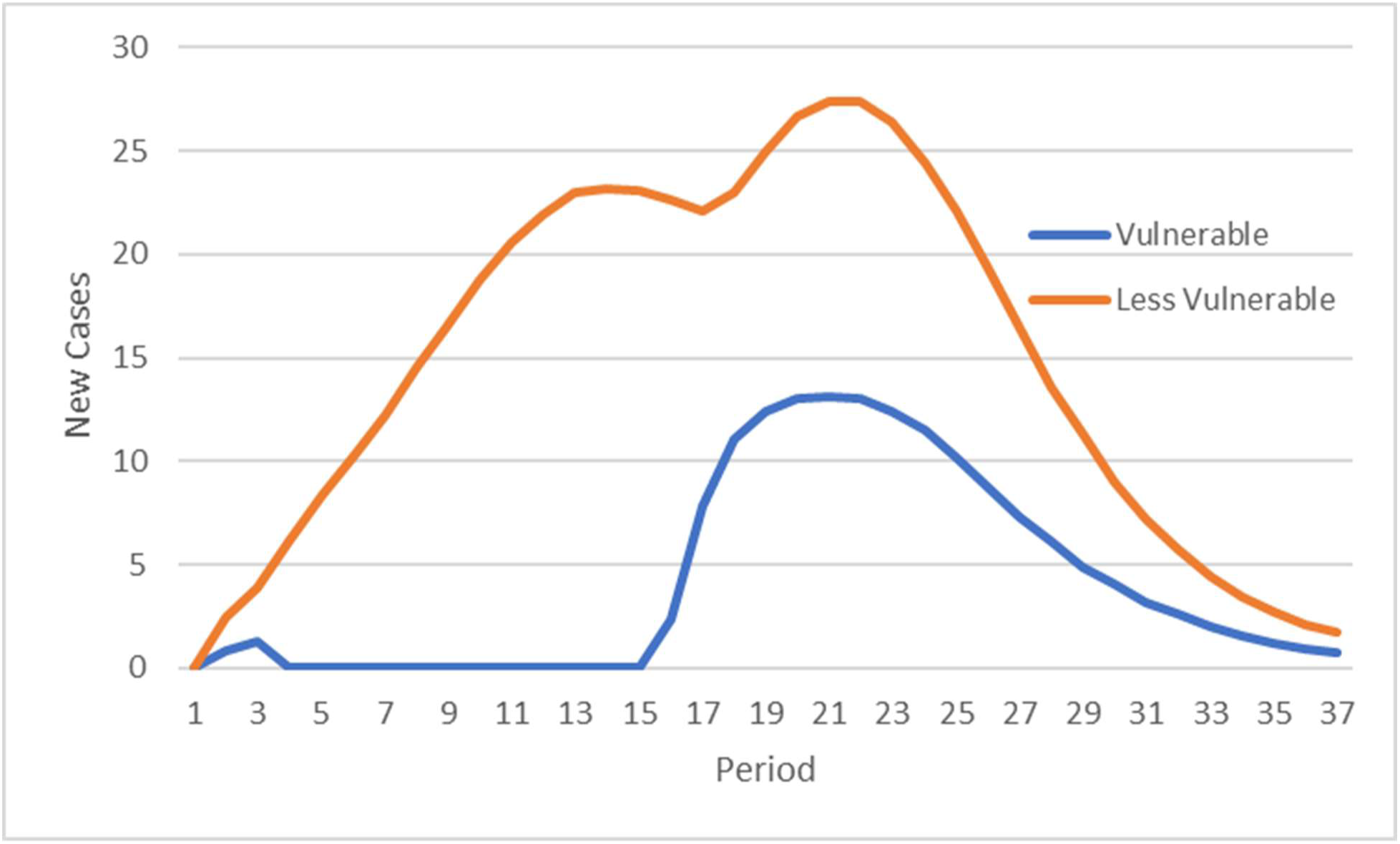
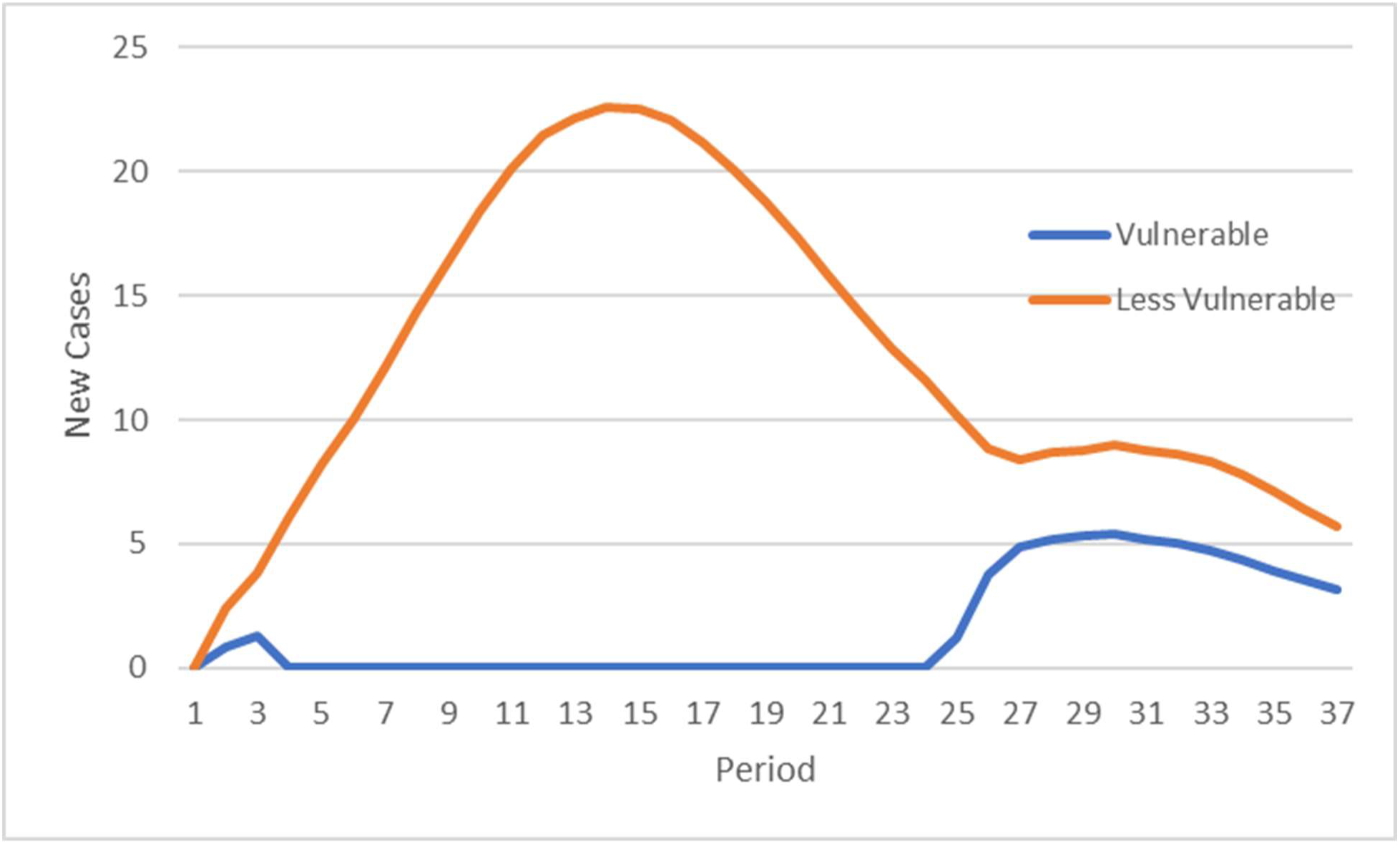
Spread of disease through populations with identical characteristics, classifying 25% of individuals as vulnerable to worse disease outcomes. A) Baseline model with simple interaction pattern (most contacts with core group, 10% with rest of population). B) Vulnerable individuals have 99% reduced contacts from period 4 to 15. C) Vulnerable individuals have 99% reduced contacts from period 4 to 24.

## Discussion

Simulation can provide a useful tool for understanding disease spread and the impact of potential interventions, both for a population as a whole and for specific defined subgroups of interest. In this paper, we built a simple model that provides a useful proxy for the spread of disease through the US population. While the stages of the model are much shorter than they would be in a larger population, interactions between individuals lead to spread of disease in patterns that are generally similar to how disease behaves in the US population. Populations where interaction networks are broader (urban populations and essential workers in this model) see faster spread of disease and can exacerbate spread in other populations.

Interventions can provide temporary or more permanent declines in the infected population. Ideally, early efforts can reduce the number of infected individuals and identify/isolate remaining cases or those newly introduced from exogenous sources. Societies where it is possible to maintain the extent and time of isolation until there are no more active cases can eliminate disease (Figure 5), but this level of control is very challenging in the US; proposals for virtually complete shut downs in the US, which could eliminate all endogenous cases, are opposed by approximately 36% of the population ^23^. However, in the absence of vaccines, disease is likely to reemerge and development of herd immunity through careful balance is difficult ^24^.

Vaccination can reduce disease spread both by protecting specific individuals and by reducing the number of interactions that spread disease (herd immunity). However, if vaccines are not completely effective or widely used, they will not fully contain the disease. A vaccine that is 50% effective would reduce effective R_0_ from about 2.5 to 1.25 if used by essentially the entire population; this would slow, but not halt the spread of the disease. However, many might refuse to take a vaccine, and it is unclear that a vaccine–or previous infection—will provide long-lasting protection. Estimates of 43% overall vaccine efficacy (combination of utilization and efficacy) in a very heterogenous population ^10^ or 60% or higher efficacy in a general population ^25^ have been suggested to provide substantial benefit in line with what would be necessary to reduce effective R_0_ sufficiently to minimize disease spread, generally in line with the above modeling.

Ultimately, it is likely that a variety of methods will be necessary to reduce the extent and spread of COVID-19 and reach the best achievable outcomes. Short-term plans to control spread need to serve feasible longer-term objectives; they will not lead to substantial reductions in the overall number of disease cases by themselves unless they virtually eliminate all disease reservoirs. If they can provide a bridge to highly effective vaccines, they can reduce overall morbidity and mortality. If the first generation of vaccines are not sufficiently effective, it is important to consider some combination of shorter-term extreme isolation, development of vaccine and natural immunity in less vulnerable populations, and heightened disease tracking/quarantine of local outbreaks.

## Limitations

Simulations and modeling are highly dependent on the assumptions and approaches that underlie them. Assumptions in this model were guided by preliminary real COVID-19 data for both inputs and reasonableness testing on outputs, but this does not guarantee that the system will behave similarly to the real world when variables are perturbed; relative effects (changes from baseline) are likely to be more robust than absolute projections. Also, these parameters are preliminary and in many cases have not been peer reviewed and thus results should be interpreted cautiously. The model utilized a small population that may have exaggerated some effects and certainly impacted stage timing relative to larger groups, and the nature of cohorts (rounds of sick patients) may have impacted model behavior. Random infection from exogenous sources does not reflect the varying likelihoods of international contacts by differently situated individuals. There remains substantial uncertainty regarding key parameters for COVID-19, including how easily it is spread between individuals and whether infected individuals become (and remain) immune to reinfection over the long term. The modeling of the US population utilized convenient important subpopulations, but does not reflect the real heterogeneity of the US population, as one might more appropriately do with an agent-based model. Patients are not distinguished by severity of disease, including the large group of individuals who may be asymptomatic. Excel random number generation is imperfect and can marginally affect results.

## Data Availability

Modeling outputs referred to in the paper are available upon request.

## Appendix

Hamidi et al. ^26^ and Wheaton and Thompson ^27^ explored whether density is an important factor in the rate of spread of SARS-CoV-2 and came to opposite conclusions. Both used county level data from early in the pandemic that may be influenced by the non-uniform distribution of early cases and challenges created (as noted by Hamidi et al. ^26^) with urban sprawl and the potential for spread to regions close to urban hotspots.

State level data can provide additional insight into this question. While not claiming to resolve this issue, an analysis of the state-level correlation of population density and number of virus-related deaths per million (See Figure A1) is supportive of the hypothesis that population density is related to viral spread. Factors like typical travel patterns (within and between states), healthcare resources, levels of comorbidities and interventions to control spread are important, but beyond the scope of this paper. While not sufficient to conclude that density is the critical factor, its use as a proxy for factors that increase the typical number of contacts for some classes of individuals is plausible.

## Financial Disclosure Summary

Daniel Mytelka is a former employee of Eli Lilly and MIT and a former shareholder of Eli Lilly. These relationships terminated before the conception of this work.

---

**Appendix Figure A1.**
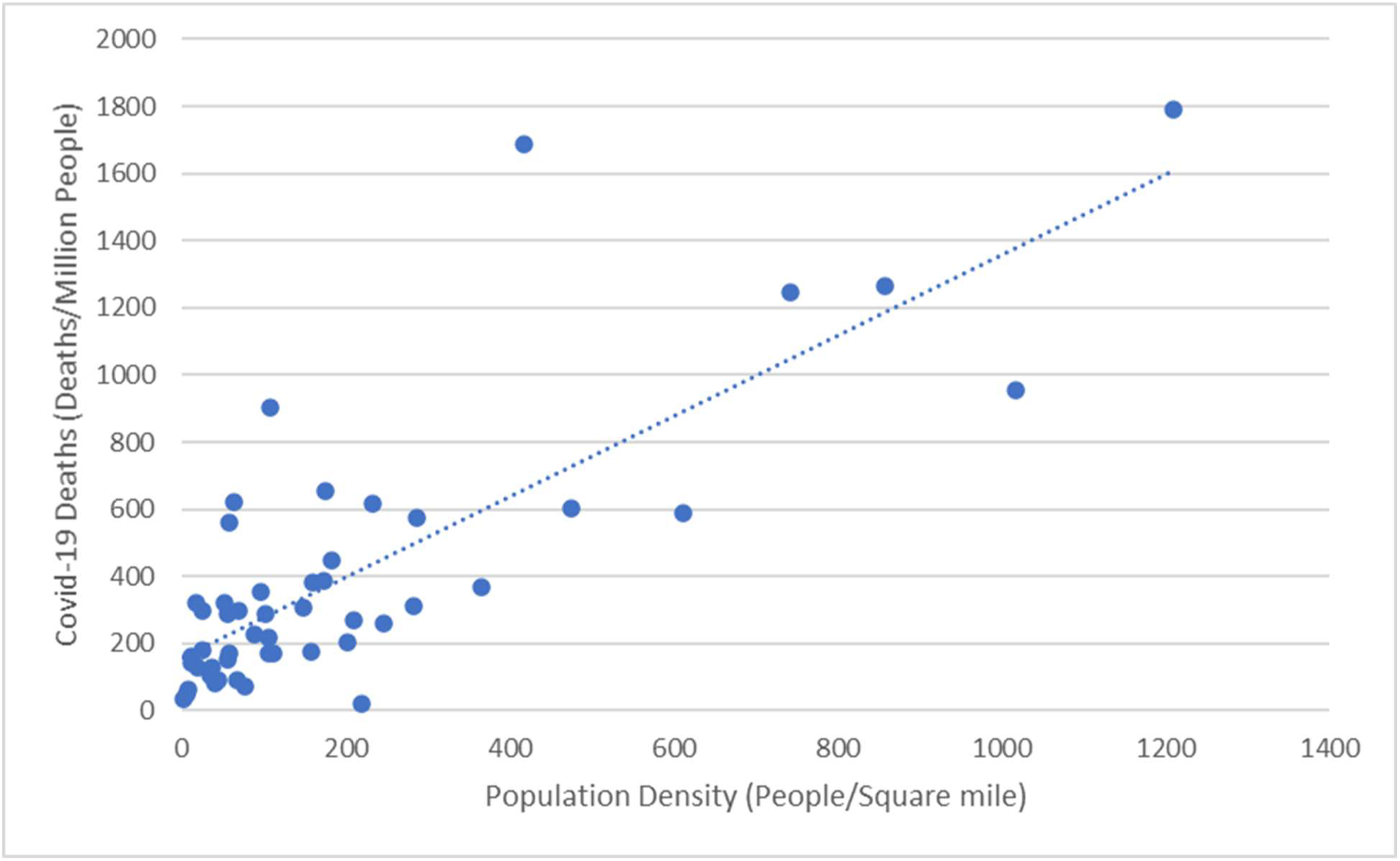
Relationship between population density ^30^ and COVID-19 deaths through August 8, 2020 ^1^ for the 50 states. R^2^ for the fit line = 0.64.

## Notes

### Funding Statement

There was no external funding for this work.

